# Vasectomy workforce and utilization in the United States, 2019

**DOI:** 10.1101/2023.04.30.23289270

**Authors:** Mandar Bodas, Julia Strasser, Rachel Banawa, Qian Luo, Ellen Schenk, Candice Chen

## Abstract

**Objective:** To examine the workforce that provides vasectomies in the United States using national-level medical claims data (IQVIA Dx, 2019).

**Methods:** We combined IQVIA Dx 2019 data on medical claims with information on clinician characteristics and clinician type from the National Plan and Provider Enumeration System (NPPES) and the American Medical Association (AMA) Masterfile. We assessed state-level trends in vasectomy provision. We used multivariate regressions to evaluate the association between clinician characteristics and two outcomes: providing at least one vasectomy and total volume of vasectomies in 2019.

**Results:** We found total of 147,618 vasectomies performed by 7,592 clinicians in the IQVIA Dx 2019 data. About 76% clinicians were urologists, 16% family medicine specialists, and about 8% were general surgeons. Urologists performed about 91% of all vasectomies. Overall, about 92% of clinicians were located in urban areas. Multivariate regression analysis showed that clinicians that were female (Odds Ratio (OR) 0.12, 95% Confidence Interval (95% CI) (0.09 - 0.15)), graduated from osteopathic schools (OR 0.77, 95% CI (0.60 - 0.98)) had lower odds of providing vasectomies.

**Conclusions:** Clinicians from multiple specialties performed vasectomies in the U.S. Most often, this procedure was performed by male urologists practicing in urban areas. Wide state-level variations exist in vasectomy provision. Clinicians’ gender, location and type of medical degree received were associated with their vasectomy provision.

## Introduction

Vasectomy is the only form of permanent contraception available to men. Mostly office-based, this effective procedure remains underutilized in the United States. A recent review of National Survey of Family Growth (NSFG) reports that the proportion of men aged 18 to 45 years that had undergone a vasectomy remained steady at around 4% between 2002 to 2017 ^1^. This is markedly lower compared to other developed Western countries, where as many as 20% of men in similar age groups use this procedure ^2^.

Research suggest that multiple factors could have potentially contributed to the stagnancy in vasectomy utilization including a sharp rise in the use of Long-Acting Reversible Contraceptives (LARCs) among reproductive age women over the past decade and the economic recovery after the 2008 mortgage crisis^3–5^. Lack of accurate knowledge may be another barrier ^6^. Research suggests that a small percentage of men receive any form of contraceptive counseling and less than 3% receive counseling on vasectomy ^7 8^. Barriers also exist due to policy issues. Vasectomies were not part of the so-called ‘contraception mandate’ of the Affordable Care Act (ACA), which meant that insurance companies did not have to cover them or other male contraceptives ^9^. While recent guidelines expanded coverage to include male condoms, coverage for vasectomy continues to be excluded. Another possible reason could be a dearth of clinicians that perform this procedure, especially in office settings; which is convenient for consumers ^10^. Previous research has found that lack of access to trained clinicians is a barrier to receiving vasectomies in publicly funded family planning organizations ^11^. Another study of Planned Parenthood clinics operating in California showed that only 5% provided vasectomy services ^12^.

Despite the known barriers around clinician availability, not many studies have analyzed vasectomy clinicians ^13,14^. Existing literature shows that about 500,000 vasectomies are performed in United States each year ^15,16^. While more than 80% of vasectomies are performed by urologists, family practice physicians and general surgeons also perform this procedure. Yet, much remains unknown about the overall workforce that provides vasectomies in the United States, including the geographic distribution of vasectomy clinicians across the country. Further, it is not fully known whether clinician characteristics such as their age, gender and the type of medical degree received (Doctor of Allopathic Medicine (MD) or Doctor of Osteopathy (DO)) influence their likelihood of performing vasectomies ^3,13,17^. Using comprehensive national-level claims data, we aim to fill this gap in the literature. As a unique contribution, we also provide state-level estimates of vasectomy provision, which are not currently available due to data limitations. Prior reports provide estimates for United States geographic regions but not for individual states. By analyzing the vasectomy workforce, we hope to uncover factors associated with a known barrier to accessing an important contraceptive method.

## Methods

### Data

The primary data source for this study was the 2019 IQVIA pre-adjudicated commercial medical claims data set (IQVIA Dx)^18^. It includes an estimated 84% of all physicians registered with the American Medical Association (AMA). IQVIA Dx data includes claims for all payer types. We obtained full-year 2019 clinician-month level counts for the procedure (CPT) code associated with contraceptive services, including vasectomy surgery (55250). We used the National Provider Identifiers (NPI) available in this data and merged it with the National Plan and Provider Enumeration System (NPPES) dataset (2020) to identify clinician type and location (state and county) of all active clinicians^19^. Further, we used the AMA Masterfile data to measure the number of years since obtaining medical degree and the type of degree received.

### Sample

Since three specialties performed more than 99% of vasectomies in our data (urology, general surgery and family medicine), we focused on a subset of clinicians from these specialties for all our analysis. For multivariate analyses, we excluded those clinicians for whom comprehensive medical claims were not available due to coverage limitations of the IQVIA Dx data and those for whom demographic information was missing. After applying these restrictions, the sample for multivariate analyses was 23,304 clinicians from family medicine, general surgery and urology specialties. A total of 3,185 clinicians in our sample performed vasectomies in 2019. Supplementary Figure 1 describes the sample derivation process.

### Measures

The primary outcome measure of our analysis was: performing at least one vasectomy in 2019. The second outcome measure was the volume of vasectomies performed in 2019. In multivariate analysis, we controlled for several clinician-level characteristics based on prior research and by specialty (urology, family medicine or general surgery), location in urban areas, type of medical degree received (MD or DO), years since graduation (categorized as 1 to 10, 11 to 20, 21 to 30, and more than 30 years), and geographic region (South, West, Midwest and Northeast).

### Analysis

We began with a descriptive analysis of the 2019 IQVIA Dx and other merged datasets. First, we identified all clinicians who performed vasectomies using CPT codes for this procedure (55250) and created a clinician-level data that included their yearly vasectomy volumes ^20^. We then merged this data with the NPPES and determined clinicians’ locations in 50 contiguous states and in Washington, DC. We excluded clinicians in US territories. Monthly volume of vasectomies at the national-level along with state-level trends in vasectomy provision were calculated as part of the descriptive analysis.

We used multivariate logistic regression models to understand the relationship between clinician characteristics and the probability of providing at least one vasectomy. Provider characteristics were selected based on prior research about clinicians’ service provision. Multivariate negative binomial regression models were used to evaluate the association between clinician characteristics and the volume of vasectomies performed in 2019. Since urologists performed the majority of vasectomies, we conducted separate analysis which focused on them. Stata/MP 17 was used to conduct all analyses ^21^. This study was approved by the George Washington University Institutional Review Board.

## Results

### Descriptive results

A total of 147,618 vasectomies were performed by 7,592 clinicians in 2019 in the IQVIA Dx data (Supplementary Table 1). Across all specialties, clinicians were mostly men and practiced in urban areas. About 91% vasectomies were performed by urologists, 5.7% by family medicine, and about 3.2% by general surgeons. The Southern U.S. had the highest proportion of vasectomy clinicians followed by the Midwest, Western, and Northeast regions.

Table 1 includes state-specific vasectomy volumes and the number of vasectomy-providing clinicians. In the IQVIA 2019 Dx data, California had the highest number of vasectomy-providing clinicians (708) followed by New York (433) and Texas (430). Despite having less than half as many vasectomy clinicians (296) as California, the state in which the greatest number of vasectomies were performed was Ohio (11,4321), followed by Texas (9,688), with California as a distant third (8,242).

**Table 1:**
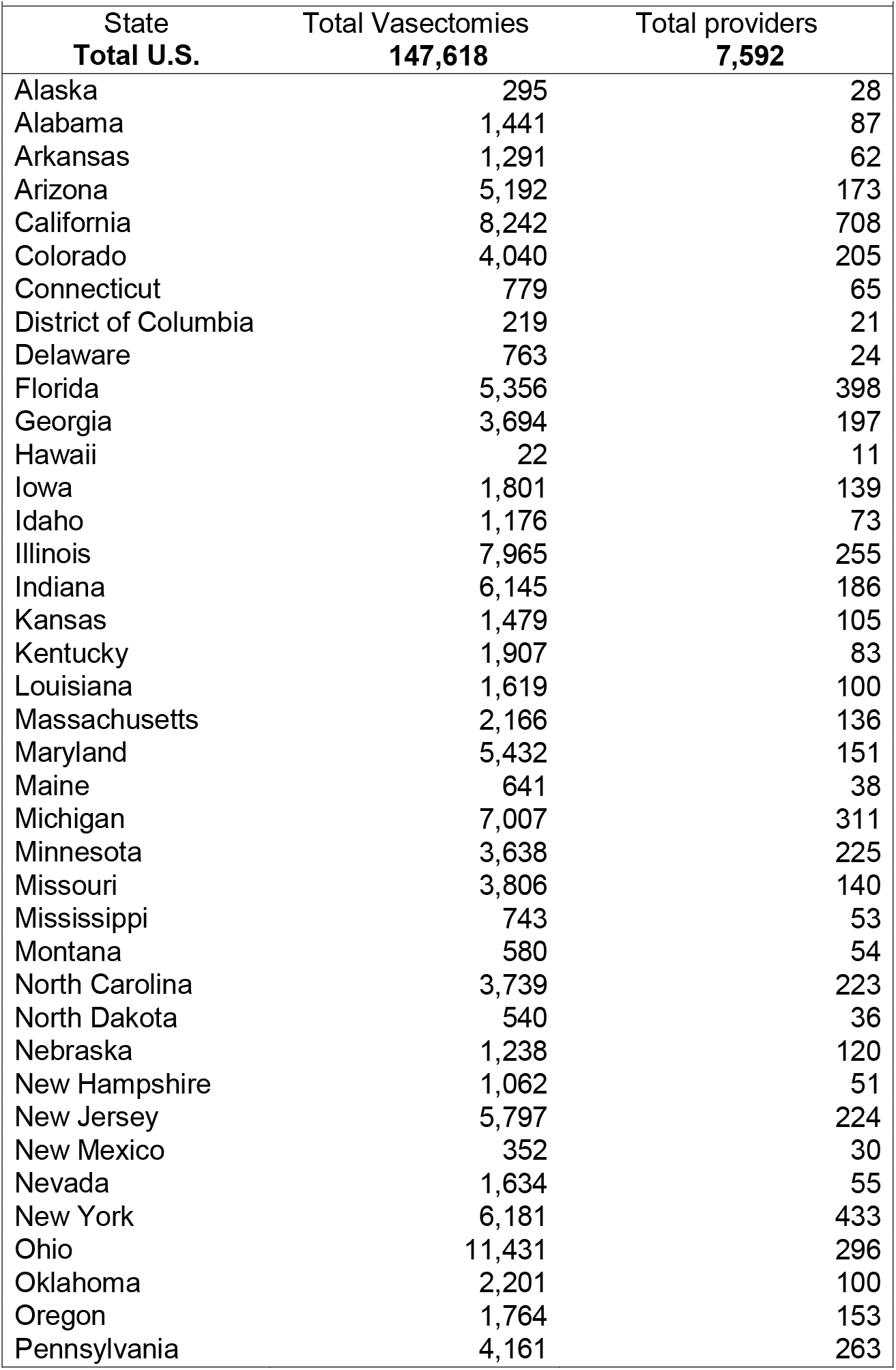
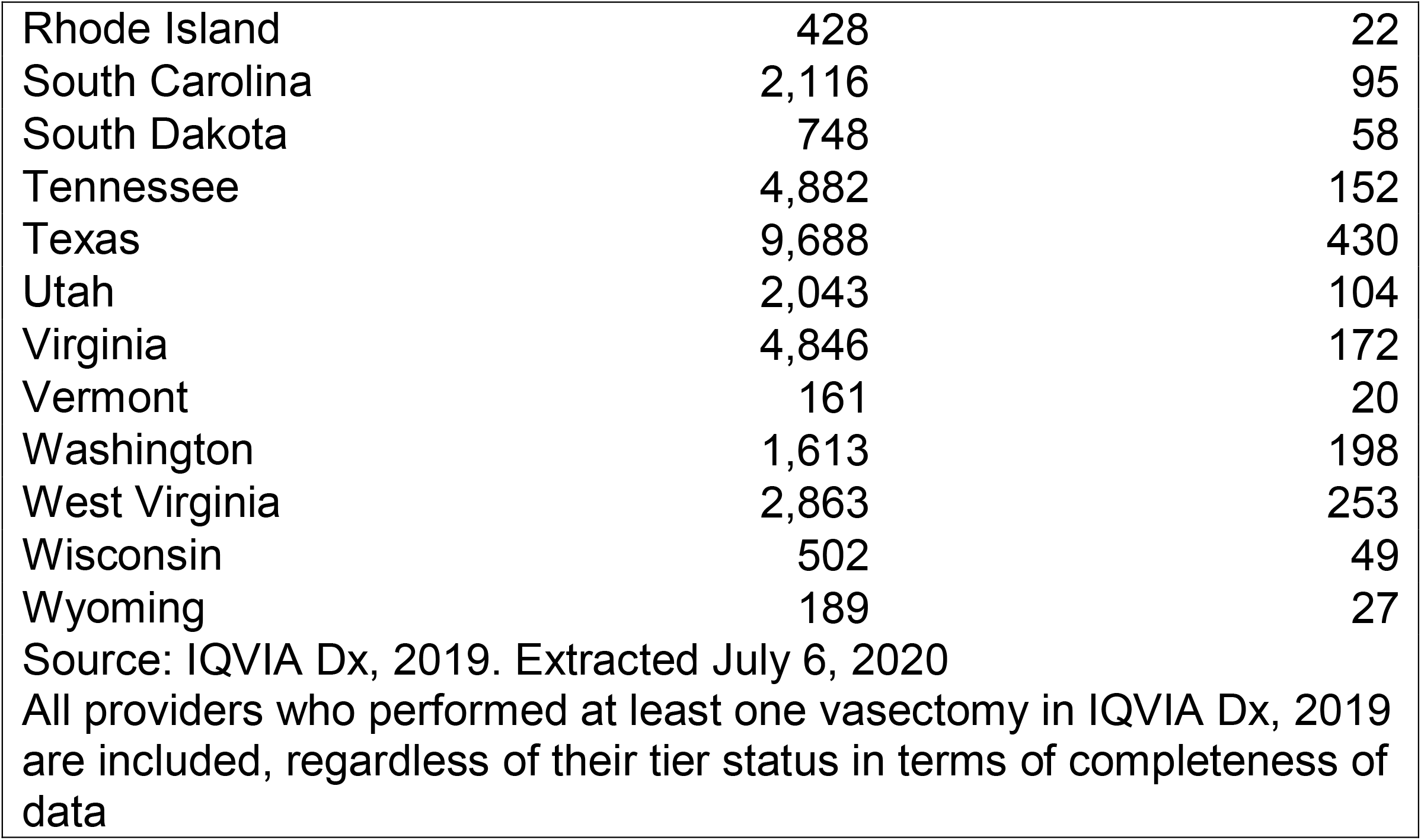
Total vasectomies and number of providers by state, 2019, IQVIA Dx

|  |  |  |
| --- | --- | --- |
| State | Total Vasectomies | Total providers |
| **Total U.S.** | **147,618** | **7,592** |
| Alaska | 295 | 28 |
| Alabama | 1,441 | 87 |
| Arkansas | 1,291 | 62 |
| Arizona | 5,192 | 173 |
| California | 8,242 | 708 |
| Colorado | 4,040 | 205 |
| Connecticut | 779 | 65 |
| District of Columbia | 219 | 21 |
| Delaware | 763 | 24 |
| Florida | 5,356 | 398 |
| Georgia | 3,694 | 197 |
| Hawaii | 22 | 11 |
| Iowa | 1,801 | 139 |
| Idaho | 1,176 | 73 |
| Illinois | 7,965 | 255 |
| Indiana | 6,145 | 186 |
| Kansas | 1,479 | 105 |
| Kentucky | 1,907 | 83 |
| Louisiana | 1,619 | 100 |
| Massachusetts | 2,166 | 136 |
| Maryland | 5,432 | 151 |
| Maine | 641 | 38 |
| Michigan | 7,007 | 311 |
| Minnesota | 3,638 | 225 |
| Missouri | 3,806 | 140 |
| Mississippi | 743 | 53 |
| Montana | 580 | 54 |
| North Carolina | 3,739 | 223 |
| North Dakota | 540 | 36 |
| Nebraska | 1,238 | 120 |
| New Hampshire | 1,062 | 51 |
| New Jersey | 5,797 | 224 |
| New Mexico | 352 | 30 |
| Nevada | 1,634 | 55 |
| New York | 6,181 | 433 |
| Ohio | 11,431 | 296 |
| Oklahoma | 2,201 | 100 |
| Oregon | 1,764 | 153 |
| Pennsylvania | 4,161 | 263 |
| Rhode Island | 428 | 22 |
| South Carolina | 2,116 | 95 |
| South Dakota | 748 | 58 |
| Tennessee | 4,882 | 152 |
| Texas | 9,688 | 430 |
| Utah | 2,043 | 104 |
| Virginia | 4,846 | 172 |
| Vermont | 161 | 20 |
| Washington | 1,613 | 198 |
| West Virginia | 2,863 | 253 |
| Wisconsin | 502 | 49 |
| Wyoming | 189 | 27 |
| Source: IQVIA Dx, 2019. Extracted July 6, 2020 |  |  |
| All providers who performed at least one vasectomy in IQVIA Dx, 2019 are included, regardless of their tier status in terms of completeness of data |  |  |

Supplementary Figure 2 shows total number of vasectomies performed each month. Consistent with prior literature, along with higher volumes in the months of November and December, we observed an upsurge in vasectomies in March^3^.

Table 2 describes the sample for multivariate analysis. As mentioned earlier, we used data on only those clinicians for which comprehensive claims information was available in IQVIA Dx dataset. The mean number of vasectomies performed was much higher among vasectomy-providing urologists (39) compared to other specialties (family medicine (8) or general surgeons (10)). Slightly over half of family medicine practitioners who performed vasectomies practiced in the Midwest (51%) and a similar trend was observed among general surgeons (42%). Additional details about the sample are available in Supplementary Table 2.

**Table 2:**
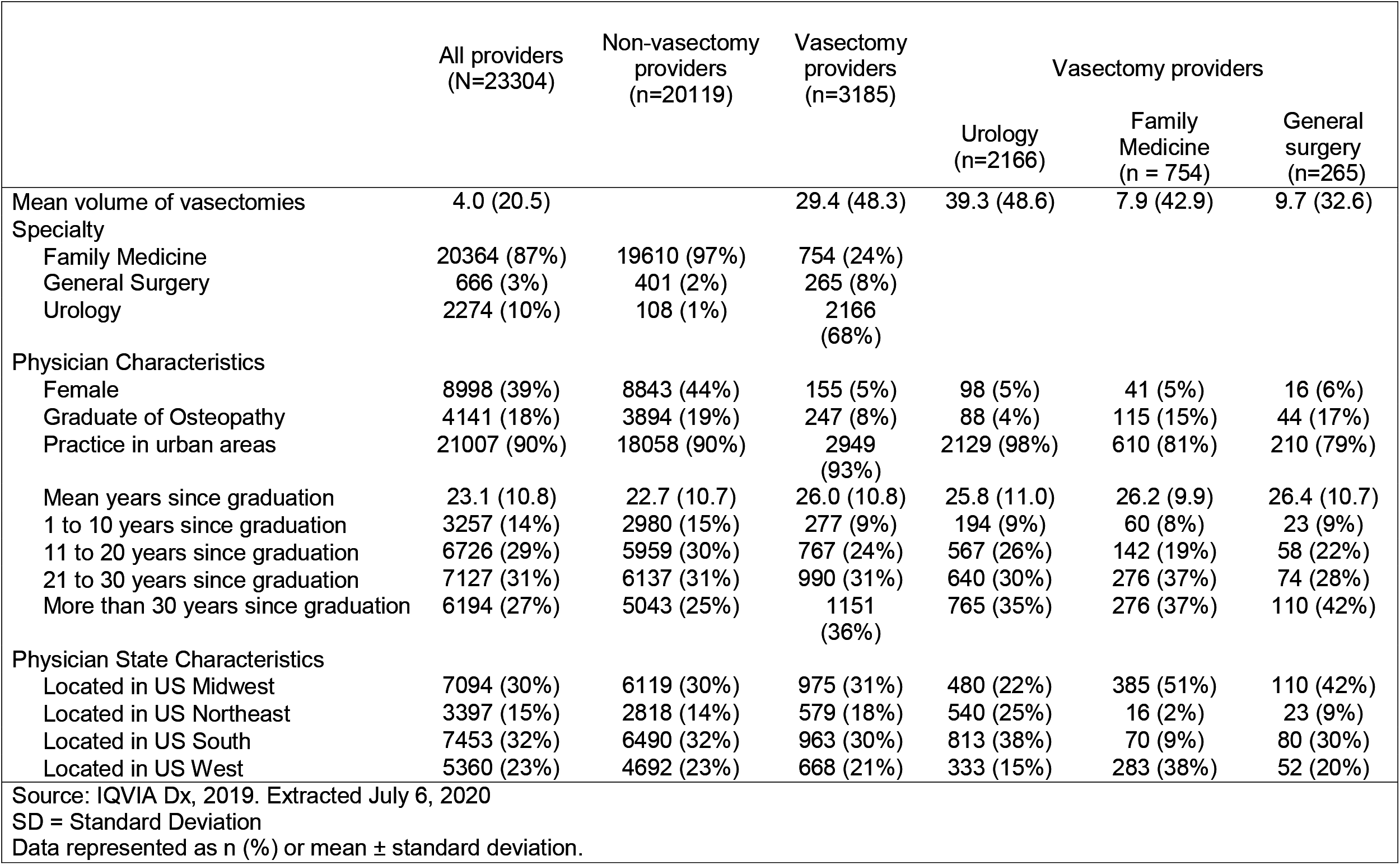
Characteristics of physicians in the analytical sample, IQVIA Dx, 2019 (N=23,304)

### Results from multivariate regression analysis

Table 3 contains the results from multivariate analysis for providing at least one vasectomy for the full sample. Compared to male clinicians, female clinicians had lower odds of providing vasectomy (Odds Ratio (OR) 0.12, 95% Confidence Interval (CI) (0.09 - 0.15)). Being a family medicine practitioner or a general surgeon was associated with lower odds of performing at least one vasectomy. Graduates of osteopathic medicine had lower odds of vasectomy provision (OR 0.77, CI (0.60 - 0.98)). Provider age had a varying relationship with study outcomes. Compared to those with more than 30 years of experience after graduation, physicians with 1 to 10 years since graduating from medical school (OR 0.64, CI (0.51 - 0.82)) and those with 11 to 20 years since graduating medical school had lower odds (OR 0.65, CI (0.53 - 0.79)) of providing vasectomies. But such an association was not seen for those with 21 to 30 years after graduation. Overall, compared to clinicians practicing in the Northeastern United States, clinicians in other regions had significantly higher odds of performing vasectomies.

**Table 3:**
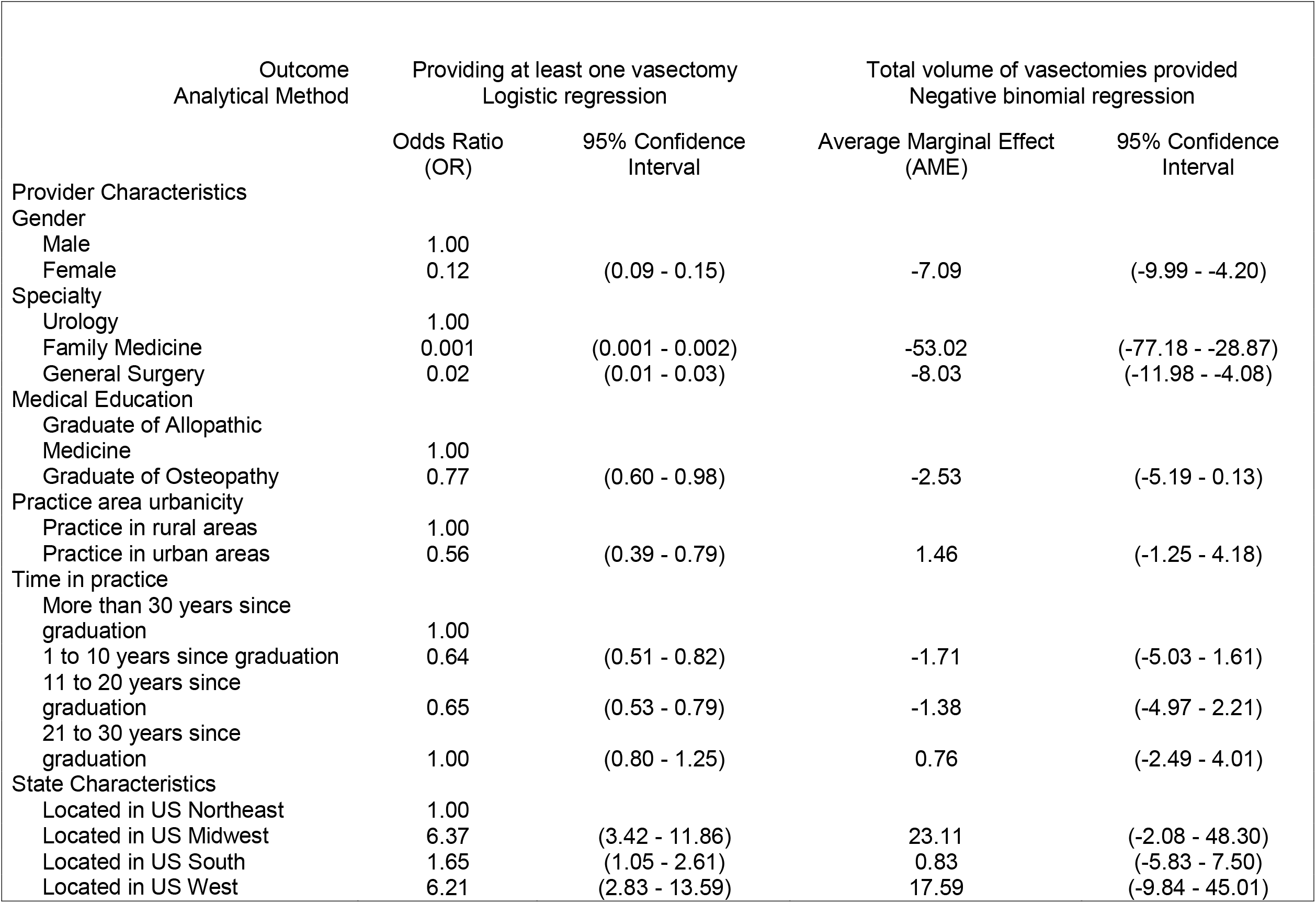
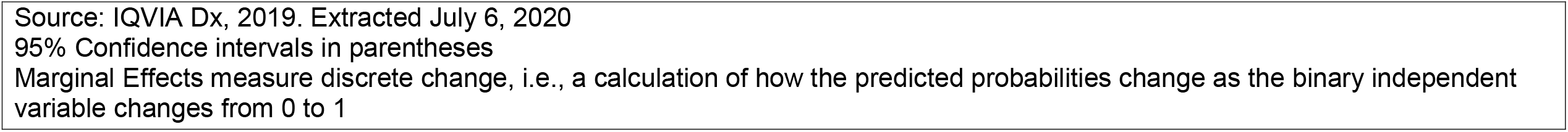
Associations between provider factors and vasectomy provision by Family Medicine Physicians, General Surgeons and Urologists in United States, IQVIA Dx, 2019 (N=23,304)

| Outcome<br>Analytical Method | Providing at least one vasectomy<br>Logistic regression |  | Total volume of vasectomies provided<br>Negative binomial regression |  |
| --- | --- | --- | --- | --- |
|  | Odds Ratio<br>(OR) | 95% Confidence<br>Interval | Average Marginal Effect<br>(AME) | 95% Confidence<br>Interval |
| Provider Characteristics |  |  |  |  |
| Gender |  |  |  |  |
| Male | 1.00 |  |  |  |
| Female | 0.12 | (0.09 - 0.15) | -7.09 | (-9.99 - -4.20) |
| Specialty |  |  |  |  |
| Urology | 1.00 |  |  |  |
| Family Medicine | 0.001 | (0.001 - 0.002) | -53.02 | (-77.18 - -28.87) |
| General Surgery | 0.02 | (0.01 - 0.03) | -8.03 | (-11.98 - -4.08) |
| Medical Education |  |  |  |  |
| Graduate of Allopathic<br>Medicine | 1.00 |  |  |  |
| Graduate of Osteopathy | 0.77 | (0.60 - 0.98) | -2.53 | (-5.19 - 0.13) |
| Practice area urbanicity |  |  |  |  |
| Practice in rural areas | 1.00 |  |  |  |
| Practice in urban areas | 0.56 | (0.39 - 0.79) | 1.46 | (-1.25 - 4.18) |
| Time in practice |  |  |  |  |
| More than 30 years since<br>graduation | 1.00 |  |  |  |
| 1 to 10 years since graduation | 0.64 | (0.51 - 0.82) | -1.71 | (-5.03 - 1.61) |
| 11 to 20 years since<br>graduation | 0.65 | (0.53 - 0.79) | -1.38 | (-4.97 - 2.21) |
| 21 to 30 years since<br>graduation | 1.00 | (0.80 - 1.25) | 0.76 | (-2.49 - 4.01) |
| State Characteristics |  |  |  |  |
| Located in US Northeast | 1.00 |  |  |  |
| Located in US Midwest | 6.37 | (3.42 - 11.86) | 23.11 | (-2.08 - 48.30) |
| Located in US South | 1.65 | (1.05 - 2.61) | 0.83 | (-5.83 - 7.50) |
| Located in US West | 6.21 | (2.83 - 13.59) | 17.59 | (-9.84 - 45.01) |

Source: IQVIA Dx, 2019. Extracted July 6, 2020
95% Confidence intervals in parentheses
Marginal Effects measure discrete change, i.e., a calculation of how the predicted probabilities change as the binary independent variable changes from 0 to 1

Similarly, Table 4 contains the results from multivariate analyses for urologists. Female urologists had lower odds of providing vasectomies (OR 0.21, CI (0.11 - 0.38)) and performed about 22 fewer vasectomies (CI, (−27.18 - -15.97)) compared to their male counterparts. Urologists with a DO degree performed about 8 fewer vasectomies (CI, (−13.94 - -1.43)). Compared to urologists who graduated from medical school more than 30 years ago, all groups of younger urologists performed more vasectomies in 2019. Similar to the findings for the full sample, compared to urologists from Northeastern United Sates, urologists from all other regions performed higher number of vasectomies, on average.

**Table 4:**
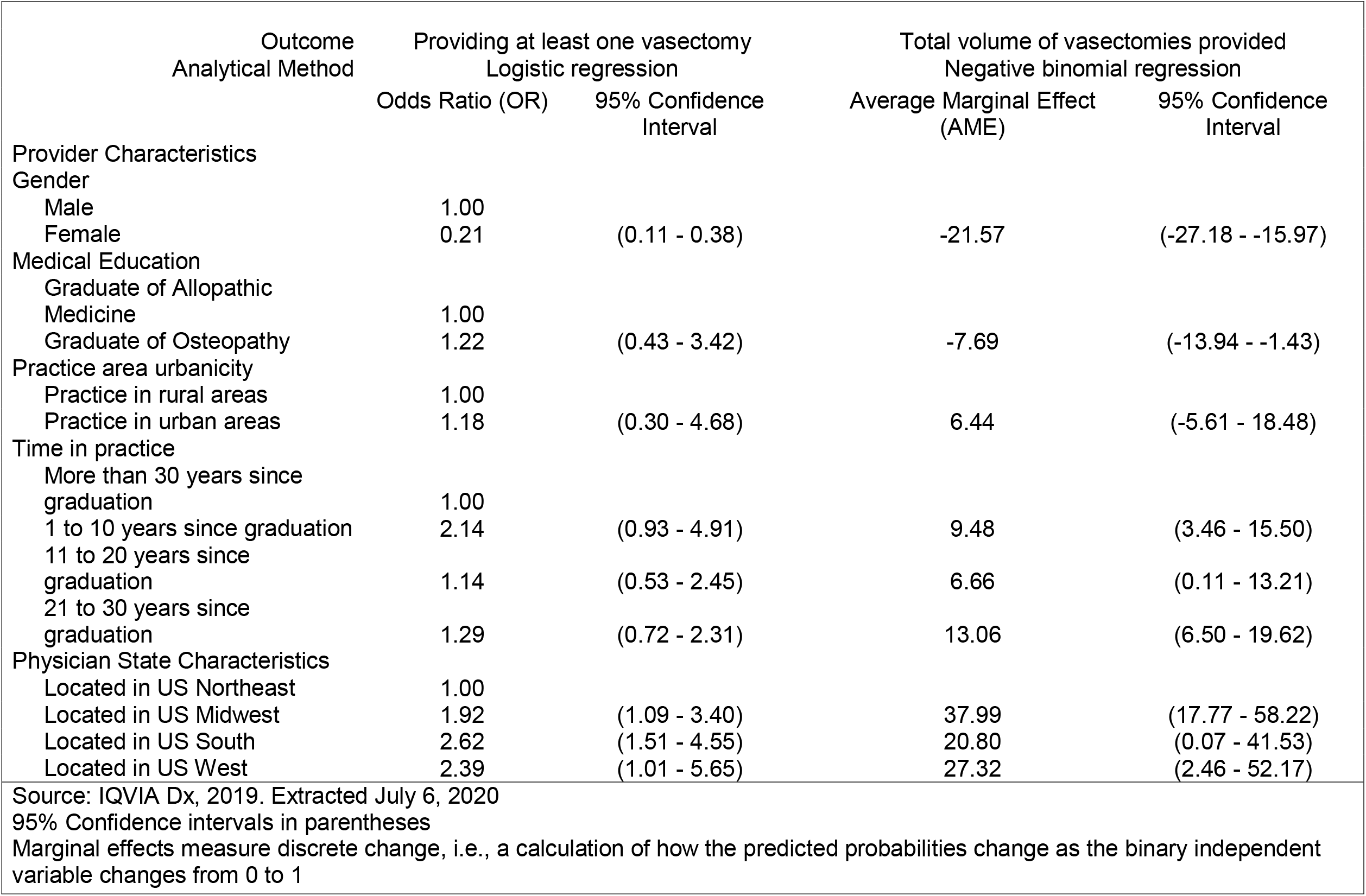
Associations between provider factors and vasectomy provision by Urologists in United States, IQVIA Dx, 2019 (N=2,274)

#### Comment

In this study, we evaluated the health workforce providing vasectomies in the United States using a comprehensive, national-level medical claims data set. Similar to prior studies, results from our descriptive analysis showed that urologists performed the greatest share of vasectomies in 2019.

The mean number of vasectomies performed by urologists in our sample was 39.2. There were about 13,044 active urologists in in 2019 according to figures released by the American Urological Association (AUA) ^22^. Assuming that urologists that were not part of our sample also perform the same number of vasectomies as our sample mean, the total number of vasectomies in the U.S. can be estimated to be about 511,325 in the year 2016. This is consistent with prior reports which suggest that nearly 500,000 vasectomies are performed in the county each year ^15,16^

Our findings about female clinicians having lower odds of providing at least one vasectomy are consistent with the evidence of a gender distribution pattern among urologists, whereby female urologists operate on a higher proportion of women than their male colleagues ^23^. This in turn, may be linked to a preference for same-gender clinicians among patients with urological problems ^24^. While exploring these and other gender-based differences (such as wages and career trajectories) among urologists is beyond the scope of this study; recent literature on the urology workforce shows that the proportion of female urologists has grown steadily in recent years ^25–27^ and nearly 25% of urologists are projected to be women in the near future ^28^. Researchers should explore how these factors will impact vasectomy provision and utilization.

Urologists with a DO degree had lower average volume of vasectomies, as seen in our multivariate analysis. While osteopathic programs started participating in ACGME-accredited urology residency match from 2016, urology was and remains a highly competitive specialty to enter for DO graduates ^29^. Future research should continue to track the presence of DOs in this workforce to determine the implications for vasectomy utilization.

We recommend that our findings at the state-level should be interpreted with caution. For instance, more vasectomies were recorded in a less populous state like Ohio compared to larger states such as Texas and California. In addition to local clinician practice patterns, demographic characteristics and patient preferences, these results could be driven by geographic coverage limitations of our data: IQVIA Dx data covers about 10% more clinicians practicing in Ohio compared to those practicing in California.

Trends observed in the monthly volume of vasectomies are consistent with prior evidence. Researchers suggest that a rise in vasectomies towards the end of the year could be attributed to patients reaching health insurance deductibles during the year and having time off work due to holidays ^15^. On the other hand, anecdotal reports suggest that relatively higher volumes in March may be related to the timing of recovery coinciding with televised programming of an athletic event - March Madness, which has high viewership among this patient population (men between the ages of 20 to 50 years) ^15^. Multiple anecdotal sources have noted this correlation, although no casual relationships have been established ^30,31^. Data from more than one year is required to confirm this trend, and there could be other factors responsible for the March vasectomy uptick such as schools having vacation.

An interesting set of findings from our multivariate analysis pertains to the impact of clinicians’ age, operationalized as years since graduation. In the regression model which included all clinician specialties, we found that younger clinicians had lower odds of providing at least one vasectomy. This could be due to the presence of a far higher number of family medicine specialists and general surgeons in our sample compared to urologists. As seen in the findings from the analysis of urologists, while age was not associated with providing at least one vasectomy; younger urologists had a significantly higher average volume of the surgery. Policymakers and medical educators should contemplate these findings in the context of documented barriers to vasectomy training for urology residents and the shortage of urologists projected by researchers ^32,33^.

There are several limitations to our analysis. IQVIA datasets have limitations in terms of completeness despite being one of most extensive sources of identified clinician information available. IQVIA indicates that their medical claims dataset covers 84% of AMA clinicians. However, the datasets may not include all claims for each of these clinicians. Based on the comprehensiveness of their claims in the dataset, IQVIA classifies clinicians in 3 tiers. For our analysis, we excluded clinicians in the third tier for whom IQVIA had limited completeness of claims. Please see Supplement for more details. Out of the 13,044 practicing urologists in 2019, our sample included 5,790 (45%) for vasectomy volume analysis and 2,166 (17%) for multivariate analysis ^22^. As a final limitation, we acknowledge that any claims dataset can only capture services for which medical claims were submitted; it is likely that we are missing vasectomies which were paid out of pocket.

## Conclusion

Despite these limitations, our study is a comprehensive analyses of vasectomy clinicians in the US. Since we relied on medical claims data, it reflects actual service provision compared to information gathered from clinician self-reported surveys. We were also able to use clinician identifiers in the IQVIA data to merge it with other data sets which complemented claims information with clinician characteristics. Finally, ours is the first study to provide details on state-specific vasectomy provision. Policymakers should be mindful of future changes in the urology workforce and its impact on the utilization of this important contraception method.

**Figure 1:**
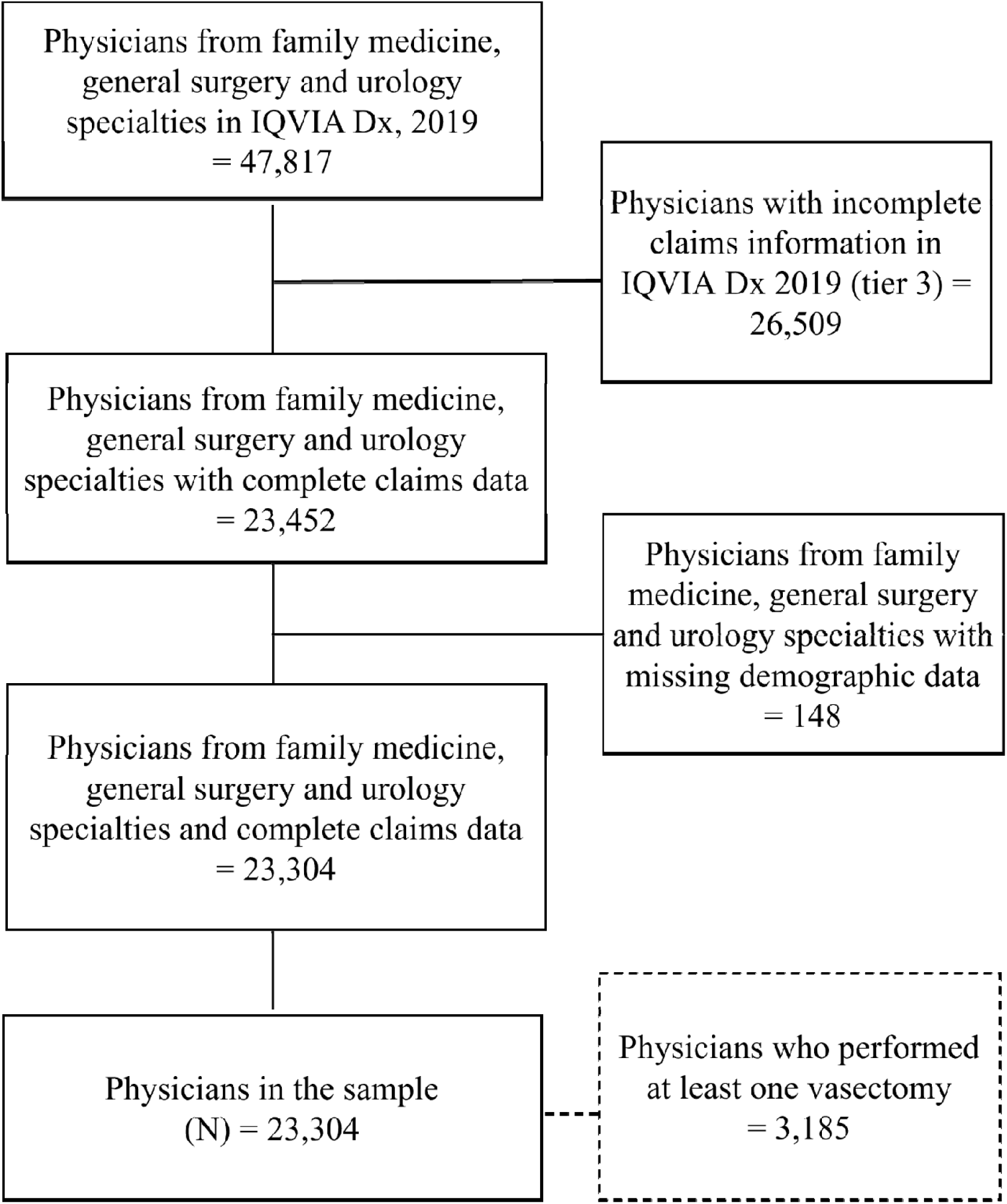
Sample Derivation Chart

## Data Availability

Data produced in this study are not available

