## Supplementary material for "Vasectomy workforce and utilization in the United States, 2019": Supplemen

Supplementary Table 1: Vasectomy providers in United States, 2019 IQVIA Dx data

Supplementary Figure 2: Monthly volume of vasectomies in United States, 2019 IQVIA Dx data

Supplementary Table 2: Vasectomy volume by month, United States, 2019 IQVIA Dx data

Supplementary Table 3: Characteristics of select physicians in the sample, 2019 IQVIA Dx data

Supplementary Figure 1: Sample Derivation Chart

Supplementary Figure 3: Additional details about study sample

IQVIA Tier Classification

| Supplementary Table 1: Vasectomy providers in United States, 2019 IQVIA Dx data, (N=7,592) | | | | |
| --- | --- | --- | --- | --- |
|  | Urology | Family Medicine | General Surgery | Overall |
| Number of providers | 5,790 (76.3%) | 1,220 (16.1%) | 582 (7.7%) | 7,592 |
| Vasectomies performed | 134,499 (91.1%) | 8,468 (5.7%) | 4,651(3.2%) | 147,618 |
| Female (%) | 7.52 | 7.38 | 8.42 | 7.57 |
| Practiced in Urban Areas (%) | 97.21 | 89.20 | 82.00 | 92.92 |
| Doctors of Osteopathy* (%) | 4.03 | 14.74 | 16.23 | 6.69 |
| Midwest (%) | 22.68 | 45.49 | 44.16 | 27.99 |
| Northeast (%) | 19.95 | 3.77 | 8.76 | 16.49 |
| South (%) | 36.74 | 9.59 | 26.29 | 31.57 |
| West (%) | 20.64 | 41.15 | 20.79 | 23.95 |
| Source: IQVIA Dx, 2019. Extracted July 6, 2020  All providers who performed at least one vasectomy in IQVIA Dx, 2019 are included, regardless of their tier status in terms of completeness of data | | | | |

Supplementary Figure 1: Monthly volume of vasectomies in United States, 2019 IQVIA Dx data

Supplementary Figure 1: Monthly vasectomy volume, IQVIA Dx 2019

|  | Vasectomies |
| --- | --- |
| January | 10952 |
| February | 10823 |
| March | 13371 |
| April | 12109 |
| May | 12514 |
| June | 10219 |
| July | 9860 |
| August | 12033 |
| September | 11166 |
| October | 12362 |
| November | 14549 |
| December | 17660 |

| Supplementary Table 2: Vasectomy volume by month, United States, 2019 IQVIA Dx data | |
| --- | --- |
| Month | Vasectomies |
| January | 10952 |
| February | 10823 |
| March | 13371 |
| April | 12109 |
| May | 12514 |
| June | 10219 |
| July | 9860 |
| August | 12033 |
| September | 11166 |
| October | 12362 |
| November | 14549 |
| December | 17660 |
| Source: IQVIA Dx, 2019. Extracted July 6, 2020  All providers who performed at least one vasectomy in IQVIA Dx, 2019 are included, regardless of their tier status in terms of completeness of data | |

| Supplementary Table 3: Characteristics of physicians in the sample, 2019 IQVIA Dx data | | | | | | | | | |
| --- | --- | --- | --- | --- | --- | --- | --- | --- | --- |
|  | All providers | Non-vasectomy providers | Vasectomy providers | All urologists | Urology vasectomy providers | All Family Medicine physicians | Family Medicine vasectomy providers | All general surgeons | General surgery vasectomy providers |
| N | 23,304 | 20,119 | 3,185 | 2,274 | 2,166 | 20,364 | 754 | 666 | 265 |
| Mean volume of vasectomies, (SD) | 4.02 (20.52) |  | 29.39 (48.34) | 37.40 (48.15) | 39.27 (48.59) | 0.29 (8.39) | 7.92 (42.93) | 3.87 (21.08) | 9.72 (32.59) |
| Specialty |  |  |  |  |  |  |  |  |  |
| Family Medicine, n (%) | 20364 (87%) | 19610 (97%) | 754 (24%) |  |  |  |  |  |  |
| General Surgery, n (%) | 666 (3%) | 401 (2%) | 265 (8%) |  |  |  |  |  |  |
| Urology, n (%) | 2274 (10%) | 108 (1%) | 2166 (68%) |  |  |  |  |  |  |
| Physician Characteristics |  |  |  |  |  |  |  |  |  |
| Female, n (%) | 8998 (39%) | 8843 (44%) | 155 (5%) | 115 (5%) | 98 (5%) | 8788 (43%) | 41 (5%) | 95 (14%) | 16 (6%) |
| Graduate of Osteopathy, n (%) | 4141 (18%) | 3894 (19%) | 247 (8%) | 92 (4%) | 88 (4%) | 3972 (20%) | 115 (15%) | 77 (12%) | 44 (17%) |
| Practice in urban areas, n (%) | 21007 (90%) | 18058 (90%) | 2949 (93%) | 2235 (98%) | 2129 (98%) | 18194 (89%) | 610 (81%) | 578 (87%) | 210 (79%) |
| Mean years since graduation, (SD) | 23.14 (10.80) | 22.70 (10.74) | 25.96 (10.75) | 25.86 (11.04) | 25.81 (11.05) | 22.78 (10.71) | 26.22 (9.90) | 24.86 (11.23) | 26.37 (10.67) |
| 1 to 10 years since graduation, n (%) | 3257 (14%) | 2980 (15%) | 277 (9%) | 200 (9%) | 194 (9%) | 2993 (15%) | 60 (8%) | 64 (10%) | 23 (9%) |
| 11 to 20 years since graduation, n (%) | 6726 (29%) | 5959 (30%) | 767 (24%) | 599 (26%) | 567 (26%) | 5927 (29%) | 142 (19%) | 200 (30%) | 58 (22%) |
| 21 to 30 years since graduation, n (%) | 7127 (31%) | 6137 (31%) | 990 (31%) | 669 (29%) | 640 (30%) | 6278 (31%) | 276 (37%) | 180 (27%) | 74 (28%) |
| More than 30 years since graduation, n (%) | 6194 (27%) | 5043 (25%) | 1151 (36%) | 806 (35%) | 765 (35%) | 5166 (25%) | 276 (37%) | 222 (33%) | 110 (42%) |
| Located in US Midwest, n (%) | 7094 (30%) | 6119 (30%) | 975 (31%) | 502 (22%) | 480 (22%) | 6387 (31%) | 385 (51%) | 205 (31%) | 110 (42%) |
| Located in US Northeast, n (%) | 3397 (15%) | 2818 (14%) | 579 (18%) | 586 (26%) | 540 (25%) | 2722 (13%) | 16 (2%) | 89 (13%) | 23 (9%) |
| Located in US South, n (%) | 7453 (32%) | 6490 (32%) | 963 (30%) | 840 (37%) | 813 (38%) | 6383 (31%) | 70 (9%) | 230 (35%) | 80 (30%) |
| Located in US West, n (%) | 5360 (23%) | 4692 (23%) | 668 (21%) | 346 (15%) | 333 (15%) | 4872 (24%) | 283 (38%) | 142 (21%) | 52 (20%) |
| Source: IQVIA Dx, 2019. Extracted July 6, 2020 SD = Standard Deviation | | | | | | | | | |

Supplementary Figure 1: Sample Derivation Chart

Physicians from family medicine, general surgery and urology specialties in IQVIA Dx, 2019

= 47,817

Physicians from family medicine, general surgery and urology specialties with complete claims data

= 23,452

Physicians from family medicine, general surgery and urology specialties and complete claims data

= 23,304

Physicians with incomplete claims information in IQVIA Dx 2019 (tier 3) = 26,509

Physicians from family medicine, general surgery and urology specialties with missing demographic data

= 148

Physicians in the sample

(N) = 23,304

Physicians who performed at least one vasectomy

= 3,185

Supplementary Figure 3: Additional details about study sample

Physicians from family medicine, general surgery and urology specialties in IQVIA Dx, 2019

= 47,817

Physicians practicing in family medicine, general surgery and urology specialties from tiers 1 and 2 in IQVIA Dx, 2019

= 23,452

Physicians practicing in family medicine, general surgery and urology specialties without missing demographic data and from tiers 1 and 2 of IQVIA Dx, 2019

= 23,304

Physicians from tier 3 in terms of completeness of provider claims information in IQVIA Dx, 2019

= 26,509

Physicians from family medicine, general surgery and urology specialties

that had missing demographic data

= 148

Physicians in the sample

(N) = 23,304

Physicians from tiers 1 and 2 in IQVIA Dx 2019 who performed at least one vasectomy

= 3,185

All physicians in IQVIA Dx 2019 who performed at least one vasectomy.

*(Sample for Table 1 and Appendix Figure 1)*

= 7,592

Physicians from tier 3 in terms of completeness of provider claims information in IQVIA Dx, 2019 who performed at least one vasectomy.

= 4,407

IQVIA Tier Classification

**Tier 1** providers are full volume, stable providers.

**Tier 2** are partial volume, unstable providers.

**Tier 3** includes providers with the monthly number of visits outside of the acceptable limits or with very unstable volume.

Stable refers to continuously being part of IQVIA data.
